# Healthcare workers’ high COVID-19 infection rate: the source of infections and potential for respirators and surgical masks to reduce occupational infections

**DOI:** 10.1101/2020.08.17.20176842

**Authors:** Lotta-Maria A. H. Oksanen, Enni Sanmark, Sampo A. Oksanen, Veli-Jukka Anttila, Jussi J. Paterno, Maija Lappalainen, Lasse Lehtonen, Ahmed Geneid

## Abstract

**Objective:** To analyse the work-related exposure to SARS-CoV-2 and trace the source of COVID-19 infections in tertiary hospitals’ healthcare workers in light of the used PPE and their ability to maintain social distances and follow governmental restrictions.

**Design:** Cross-sectional study

**Setting:** Tertiary hospitals in Uusimaa region, Finland

**Participants:** Of 1072 enrolled, 866 HCWs (588 nurses, 170 doctors and 108 laboratory and medical imaging nurses) from the Helsinki University Hospital completed the questionnaire by July 15^th^, 2020. The average age of participants was 42.4 years and 772 (89.0%) were women. The participants answered a detailed questionnaire of their PPE usage, ability to follow safety restrictions, exposure to COVID-19, the source of potential COVID-19 infection and both mental and physical symptoms during the first wave of COVID-19 in Finland.

**Main outcome measures:** All participants with COVID-19 symptoms were tested with either RT-PCR or antibody tests. The infections were traced and categorised based on the location and source of infection. The possibility to maintain social distance and PPE usage during exposure were analyzed.

**Results:** Of the HCWs that participated, 41 (4.7%) tested positive for SARS-CoV-2, marking a substantially higher infection rate than that of the general population (0.3%); 22 (53.6%) of infections were confirmed or likely occupational, including 7 (31.8%) from colleagues. Additionally, 5 (26.3%) of other infections were from colleagues outside the working facilities. 14 (63.6%) of occupational infections occurred while using a surgical mask. No occupational infections were found while using an FFP2/3 respirator and aerosol precautions while treating suspected or confirmed COVID-19 patients.

**Conclusions:** While treating suspected or confirmed COVID-19 patients, HCWs should wear an FFP2/3 respirator and recommended PPE. Maintaining safety distances in the workplace and controlling infections between HCWs should be priorities to ensure safe working conditions.

## INTRODUCTION

SARS-CoV-2 spreads mainly via droplets, secretions and direct contact^1^. Lately, the possibility of airborne transmission has been discussed even in the absence of aerosol-generating procedures (AGP), especially indoors^2 3^. SARS-CoV-2 appears to be more infectious than influenza, and the reproductive number (R0) has been estimated to be as high as 2.3–5.7 in the general population in various studies^1 4–6^. The infection rates of healthcare workers (HCW) varies widely from country to country, ranging from 2.2% to 44% yet exceeding that of the general population^7 8^.

### The first wave and restrictions in Finland

The COVID-19 infection reached the epidemic threshold in mid-March, and the Finnish government declared a state of emergency from March 16^th^ to June 16^th^ 2020. The primary focus was on social distancing with restrictions for travel, limitations of no more than 10 persons for public gatherings, and recommendations to avoid spending time in public places. Additionally, visitors were banned from care institutions, healthcare units, and hospitals. Also, the epicenter of the COVID-19 epidemic in Finland, the Uusimaa region, was isolated from the rest of the Finland between March 28^th^ and April 15^th^. Helsinki University Hospital (HUS) is responsible for that region’s specialised care, and its healthcare professionals are the focus of this article. By July 15^th^, Finland had 7 293 confirmed cases of COVID-19, 5223 of which were in the HUS region, with a 0.3% infection rate.^9^ In the HUS region, 794 (15.2%) workers in social and healthcare organizations were infected with COVID-19 by July 15^th^ 2020; 349 (44.0%) had occupational infection, 207 (26.1%) infections were non-occupational, and for 238 (30.0%) the source was unclear; 153 were HCWs (19.3%) working at HUS.^10–12^

### Restrictions at HUS

Due to the crisis, HUS instructed personnel with several restrictions to avoid spreading the virus in the hospital facilities; non-urgent patient contact was postponed, and personnel were instructed to 1) avoid all trips abroad, 2) avoid all gatherings and favour digital meetings and remote patient contact, 3) keep at least one meter of distance from other employees, 4) maintain good hand hygiene, 5) use the required personal protective equipment (PPE), and 6) self-isolate and get tested for COVID-19 (nasopharyngeal or oropharyngeal RT-PCR) if they experience any COVID-19-related symptoms.

### The debate over masks and respirators in COVID-19 infections

As the transmission routes of SARS-CoV-2 are still partly debated, there has been speculation about which PPE (especially masks and respirators) HCWs should use with various patient groups and during medical procedures. The World Health Organization (WHO) has aligned that HCWs working with COVID-19 patients should use masks throughout their shift and N95 or FFP2/FFP3 respirators during potential AGPs as well as in semi-intensive and intensive care units (ICU)^13^. During the first wave, HUS generally followed the WHO’s recommendations regarding PPE instructions for the ICU, but no masks were used in the staff area. Cohorted wards had similar recommendations for PPE at first but were quickly reduced to surgical mask instead of an FFP3 respirator; on March 26^th^ the instruction was loosened further as gowns were only required during close patient contact, and hair protection was no longer required. Cohorted wards followed droplet precautions whereas ICUs also followed aerosol precautions while doffing.

Differences between surgical masks and respirators like N95 and FFP2/3 have been studied during the recent decade concerning different respiratory viruses and bacteria but not with COVID-19. A meta-analysis and systematic review by Smith et al. shows that N95 respirators have a protective advantage over surgical masks in laboratory conditions but do not protect HCWs better against influenza in clinical work^14^. Other studies have not shown any difference in morbidity from influenza in HCWs using either an N95 respirator or a surgical mask^15^. However, in bacterial infections, have been observed to prevent infection better than surgical masks^15^. These previous studies did not obserwe the possibility of nonoccupational infection and used no other restrictions to reduse the non-work or colleague-related exposure.

### Purpose of the study

The purpose of this study was to analyze the work-related exposure to SARS-CoV-2 and trace the source of COVID-19 infections in tertiary hospitals’ HCWs in light of the used PPE and their ability to maintain social distances and follow governmental restrictions. The hypothesis was that usage of FFP2/3 masks prevents workplace related COVID-19 infections.

## MATERIAL AND METHODS

Out of 17 740 nurses, midwives, and doctors working at Helsinki University Hospital, 1072 (6.0%) enrolled to the study, and 866 (4.9%) answered the questionnaire completely between June 12^th^ and July 15^th^, 2020. The minimum study size of 366 was calculated by N95 respirators statistical power calculation. The inclusion criteria were 18 years of age or older; education as a practical nurse, paramedic, nurse, laboratorian, radiological nurse, midwife, or doctor; and employment at HUS between March and July, 2020. Participants filled out a detailed questionnaire with 150 questions about their common health risks (smoking, alcohol consumption, body mass index, diseases, and medications as well as risk factors for COVID-19), leisure and working conditions during the COVID-19 pandemic (including PPE, potential infection symptoms, and exposure to COVID-19), and other topics about COVID-19 and mental health. Participants’ medical history was reviewed in July 2020 for COVID-19 RT-PCR and antibody results. In Finland, all HCWs including the participants of this study who presented any COVID-19-related symptoms have been tested according to the instructions of the Finnish Institute for Health and Welfare (THL) with the standard HUSLAB RT-PCR methods^16 17^. Additionally, some HUS workers have been tested with antibodies as part of the employer’s COVID-19 strategy, and all who tested positive with the first antibody test were tested for neutralising antibodies to confirm the diagnostics^18^. Potential AGPs were listed according to the THL as intubation, extubation, resuscitation, direct laryngoscopy, bronchoscopy, upper gastrointestinal endoscopy, non-invasive ventilation, use of nebulizer, high-flow nasal oxygen, open suction of the mucus from airways, and oral and ENT surgery^19^.

The number of COVID-19 infections in HCWs was calculated and compared to the general population in the same area. The infection rate was calculated for all HCWs and each occupation separately. PPE usage was compared between infected and non-infected HCWs. All infected participants were contacted, and their answers were confirmed regarding the tracing of infection, the usage of PPE at the time of the assumed transmission, and the ability to maintain social distance at that time. The original tracing was done after the initial diagnosis by a local tracing team led by an infection doctor. The infection was categorised as occupational if there were clear infection contacts (ICs) in the workplace and no possible IC outside the workplace; infection was likely occupational if there were clear ICs in the workplace and some non-COVID-19-related contacts outside the workplace. Not all infections were traced successfully, so they were marked “unclear”.

### Statistical analysis

The statistical analysis was performed using statistical software (IBM SPSS Statistics 25, Chicago, USA). Differences between nominal variables were tested using a chi-square test or Fischer’s exact test with a P< 0.05 significance level. Odds ratios (ORs) and a 95% confidence interval (CI) were calculated using logistic regression with variance calculation. Some of the participants didn’t answer to all questions, leading to varying sample size from question to question. The number of answers per question is presented accordingly.

### Patient and public involvement

Due to the exceptional situation during the COVID-19 pandemic, no patients or public were directly involved in the development, implementation, or interpretation of this study. Our study design was established on questionnaires, RT-PCR, and antibody testing, that were aimed at HUS’ HCWs and required strong identification for enrolment.

### Ethical considerations

All procedures that involved human participants were conducted in accordance with the ethical standards of the institutional or national research committee and the 1964 Declaration of Helsinki and its later amendments or comparable ethical standards. The Ethics Committee of Helsinki University Hospital approved the study protocol (HUS/1760/2016). All responders provided written informed consent prior to their participation.

## RESULTS

### Characteristics, infection rates, and exposure among studied HUS personnel

Of the 866 HCWs who participated in the study, 820 (94.7%) said that they had followed all the restrictions imposed by the Finnish government. Characteristics of the participants are presented in Table 1.

**Table 1.**
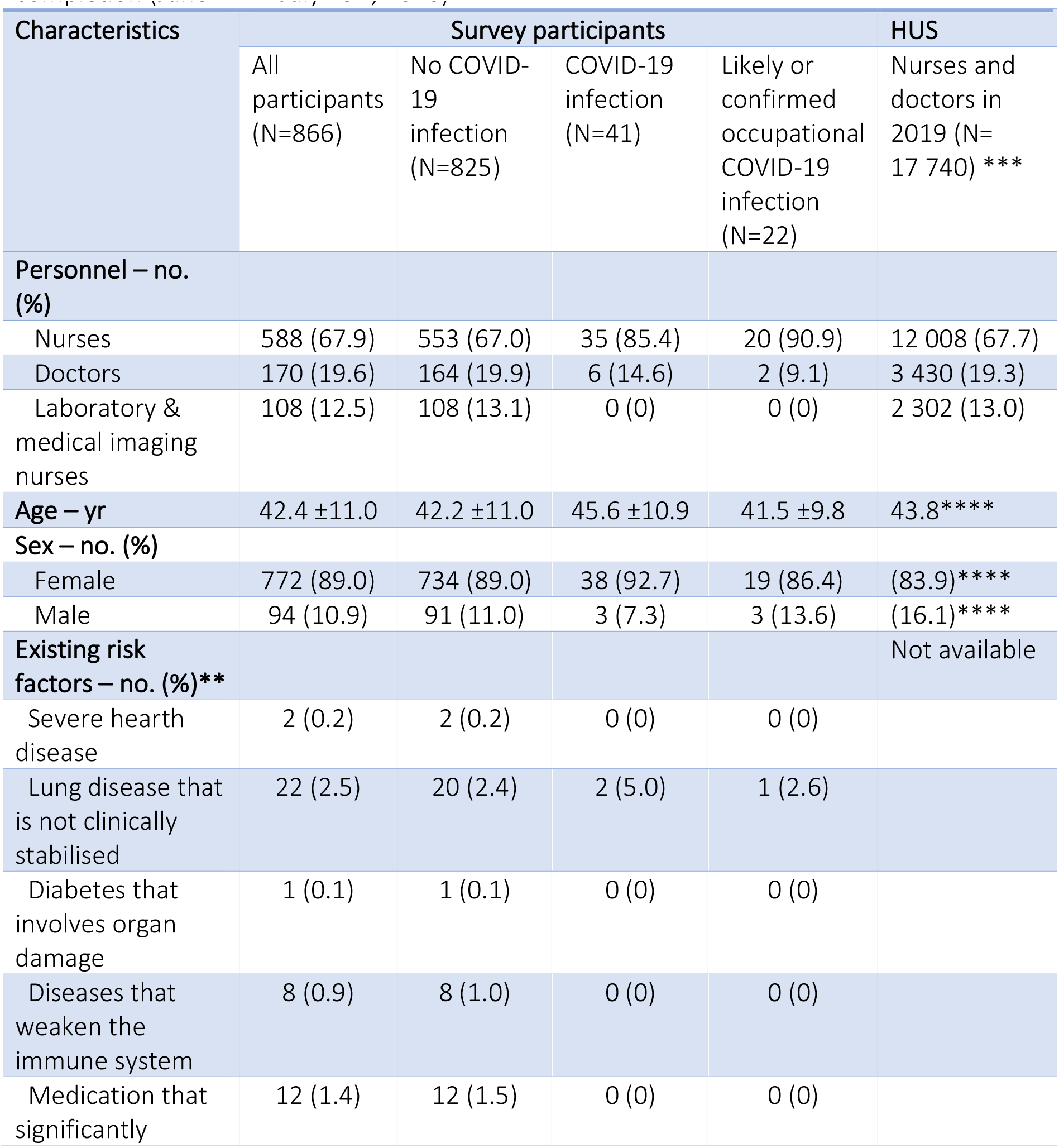
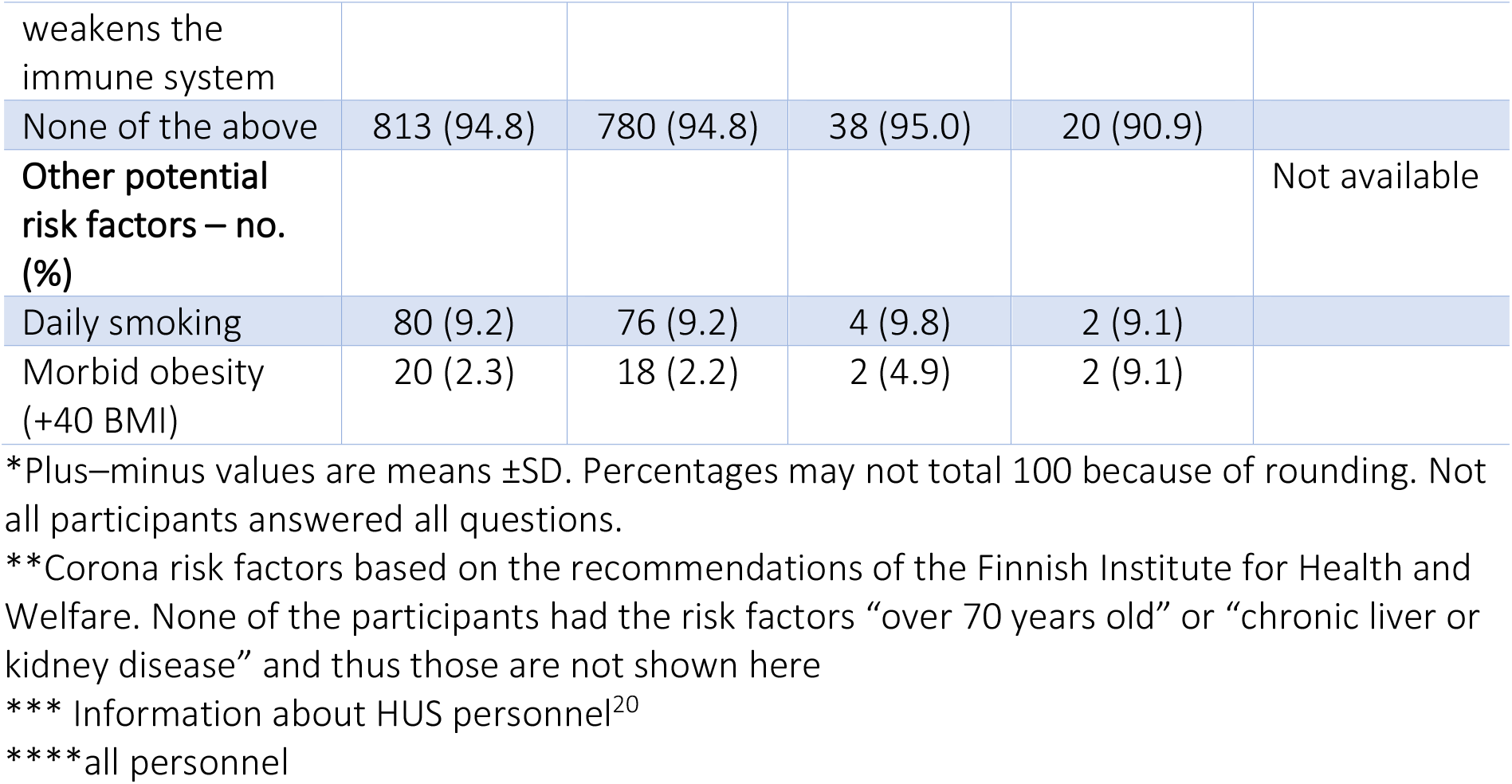
Characteristics of survey participants divided into four groups (all, no COVID-19 infection, COVID-19 infected, likely occupational COVID-19 infection) compared to personnel from HUS.* Infection status by July 15^th^, 2020 or by the time of survey completion (June 12^th^– July 15^th^, 2020)

During the first wave, 431 (52.8%) participants took care of at least some COVID-19 patients. Out of 21 pregnant participants, 10 (47.6%) knowingly took care of COVID-19 patients, and 3 (14.3%) were uncertain. Among the group, 47 (5.4%) HCWs said they had tested positive for COVID-19 either by RT-PCR or antibody test, but 6 (12.8%) were later found to be false positive as they only had a positive first antibody test and a negative neutralization test; this resulted in 41 (4.7%) true positive infections, representing 26.8% of infected HCWs at HUS. Of those who tested positive, 26 (63.4%) COVID-19-positive participants informed that they knowingly took care of COVID-19 patients.

The infection rates of COVID-19-positive participants are shown by location and source in Table 2.

**Table 2.** COVID-19 infection rate of participants (N = 866) by location of infection, and location of infection by source.

| Infection location of positive participants | Confirmed workplace (N=13) | Likely workplace (N=9) | Unclear (N=4) | Outside workplace (N=15) |
| --- | --- | --- | --- | --- |
| Infection rate by infection location, % of participants | 1.5 | 1.0 | 0.5 | 1.7 |
| Source of infection – no. (%) |  |  |  |  |
| Colleague | 5 (38.5) | 0 (0.0) | 0 (0.0) | 5 (33.3) |
| Likely colleague | 0 (0.0) | 2 (22.2) | 0 (0.0) | 0 (0.0) |
| Business trip | 0 (0.0) | 0 (0.0) | 0 (0.0) | 2 (13.3) |
| Patient | 3 (23.1) | 0 (0.0) | 0 (0.0) | 0 (0.0) |
| Likely patient | 1 (7.7) | 1 (11.1) | 0 (0.0) | 0 (0.0) |
| Unclear | 4 (30.8) | 6 (66.7) | 4 (100.0) | 1 (6.7) |
| Non-occupational | 0 (0.0) | 0 (0.0) | 0 (0.0) | 7 (46.7) |

80% of the infections origin from patients happened either in the normal or cohorted ward. Additionally, 61 (7.4%) non-infected participants were quarantined by local infection doctors due to occupational COVID-19 exposure, 13 (1.6%) due to non-occupational exposure, and 1 (0.1%) due to both. Of the 62 occupational exposures, 37 (59.7%) were from colleagues, 13 (21.0%) from patients, and 12 (19.3%) were not stated.

During the first wave, 20 (48.8%) of the infected worked either in a normal ward or among a cohort. Only 4 (20.0%) of these infected employees working in either ward were able to maintain the instructed 1-meter safety distance, whereas 8 (38.1%) of other infected were able to do so. Similarly, only one out of seven participants (16.7%) who got infection from a colleague at the workplace was able to maintain the 1-meter distance at the time of infection.

Of 93 ICU workers, no clear ICU-related infections were found. Two ICU workers reported either positive COVID-19 RT-PCR or positive antibody testing, excluding false positives. One had a non-occupational infection source and another’s remains unclear.

Further information about the workplace of participants during the first wave of the COVID-19 epidemic, ability to maintain 1-meter distance and ORs is seen in Table 3.

**Table 3.** Working place, ability to maintain 1 meter distance and OR for infection of participants during the first wave of the COVID-19 epidemic.

| Department | Was able to maintain 1-meter distance N (%)<sup>*</sup> | Infected N (%) | Any COVID-19 infection |  |  | Work-related COVID-19 infection |  |  |  |
| --- | --- | --- | --- | --- | --- | --- | --- | --- | --- |
|  |  |  | OR | %95 CI | P | OR | %95 CI | P | Combined OR (%95 CI), P |
| Cohort COVID-19 ward (N=83) | 23 (28.0) | 5 (6.0) | 1.3 | 0.5–3.5 | 0.583 | 2.9 | 1.0–8.0 | 0.051 | 3.4 (1.2–9.2)<br>0.016 |
| Ward (N=170) | 58 (34.1) | 15 (8.8) | 2.5 | 1.3–4.8 | 0.008 | 2.0 | 0.8–4.9 | 0.170 |  |
| Emergency department (N=96) | 26 (27.1) | 6 (6.3) | 1.4 | 0.6–3.4 | 0.443 | 2.4 | 0.9–6.8 | 0.086 |  |
| ICU (N=93) | 29 (30.9) | 2 (4.3) | 0.4 | 0.1–1.7 | 0.214 | n/a | n/a | 0.099 |  |
| Medical imaging (N=55) | 23 (41.8) | 0 (0.0) | n/a | n/a | 0.088 | n/a | n/a | 0.216 | 0.3 (0.1–0.8)<br>0.016 |
| Outpatient appointment clinic (N=175) | 100 (57.1) | 6 (3.4) | 0.7 | 0.3–1.6 | 0.431 | 0.2 | 0.0–1.4 | 0.064 |  |
| Researcher / administration / other non-patient (N=50) | 32 (64.0) | 3 (6.0) | 1.3 | 0.4–4.4 | 0.664 | 1.7 | 0.4–7.3 | 0.499 |  |
| Sampling (N=97) | 40 (41.2) | 2 (2.1) | 0.4 | 0.1–1.7 | 0.188 | 0.8 | 0.2–3.4 | 0.751 |  |
| Surgery (N=47) | 14 (29.8) | 2 (4.3) | 0.9 | 0.2–3.8 | 0.874 | n/a | n/a | 0.255 |  |
| All (N=866) | 345 (39.9) | 41 (4.7) | n/a | n/a | n/a | n/a | n/a | n/a | n/a |

<sup>\*</sup> One missing value.

### PPE usage of participants

Of the participants, 340 (39.5%, n = 861) practised donning and doffing (D&D) PPE under supervision, and only 77 (8.9%) answered that they did not know how to do D&D correctly. Among the participants with occupational infection, 10 (45.5%) informed that they practiced D&D under supervision, and all 22 (100%) answered that they knew how to do it correctly. Only 13 participants (1.8%) told that they used a minimum of FFP2/3 respirators with non-infection patients (Figure 1), and that figure increased to 169 (28.5%) with suspected COVID-19 patients, but even with confirmed COVID-19 patients, fewer than half (210, 41.7%) used at least an FFP2/3 respirator.

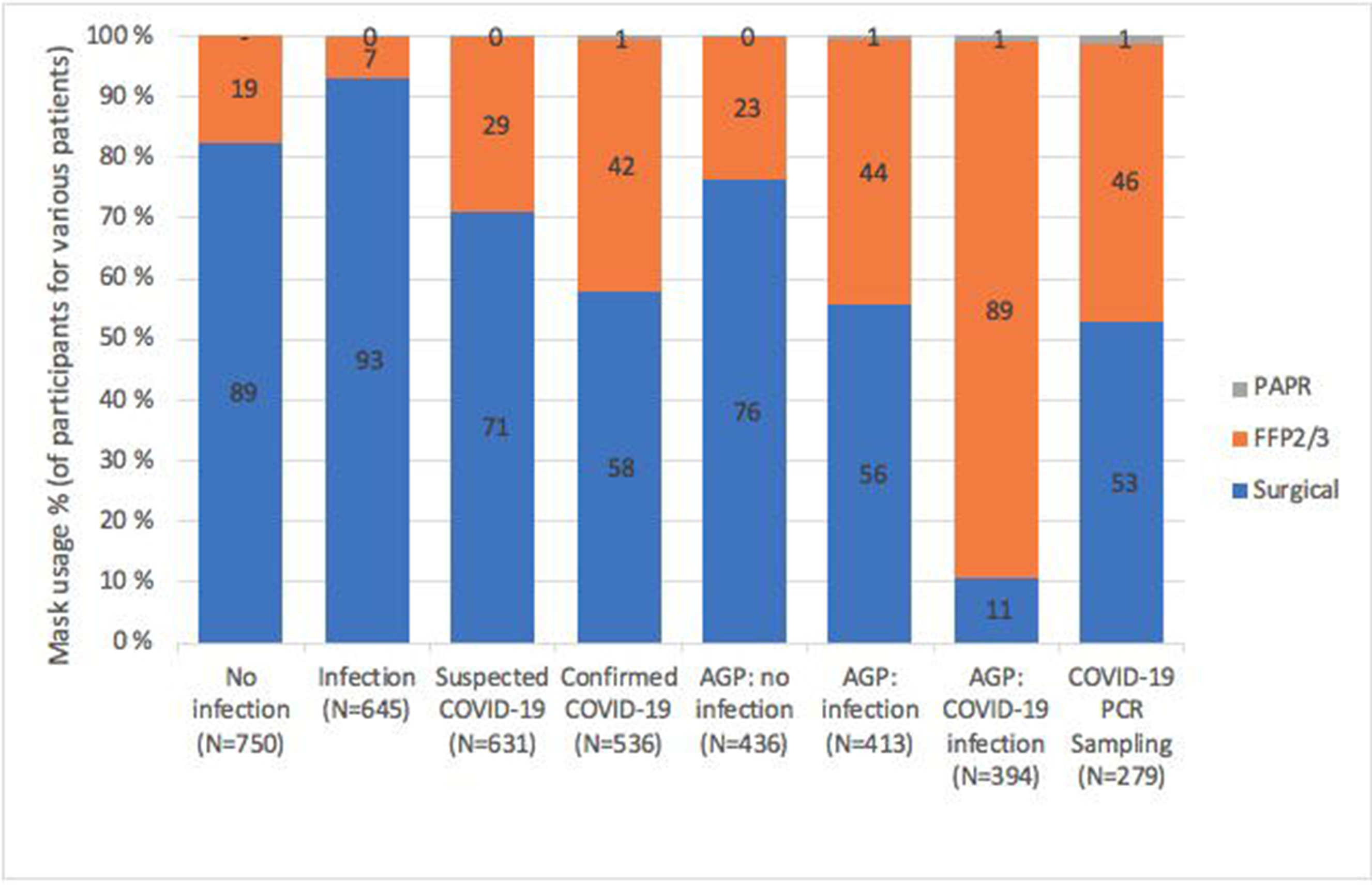

### Usage of respirators and masks, and ability to maintain social distance from COVID-19- positive participants at the time of infection

Out of 22 confirmed or likely occupational infections, only 7 (31.8%) informed that they were able to maintain a distance of at least 1 meter from other staff members. Regarding PPE shortages, 23 (2.8%, N = 824) participants reported a complete shortage of PPE, but none of them were diagnosed with COVID-19. 224 (27.2%, N = 824) other participants reported some cases of partial shortages with 7 (3.1%) confirmed or likely workplace related infections. Detailed information on positive participants’ COVID-19 infection location and mask and respirator usage is presented in Table 4.

**Table 4.** COVID-19 positive participants’ mask and respirator usage, safety distance and source of infection by infection location groups.

| Characteristics | The most probable location where infection occurred according to infection tracing |  |  |  |
| --- | --- | --- | --- | --- |
|  | Workplace (N=13) | Likely workplace (N=9) | Unclear (N=4) | Outside workplace (N=15) |
| Used mask/respirator at the estimated infection time – no. (%) |  |  |  |  |
| FFP2/3 respirator | 0 (0.0) | 0 (0.0) | 1 (25.0) | 0 (0.0) |
| Surgical mask | 9 (69.2) | 5 (55.6) | 1 (25.0) | 0 (0.0) |
| None | 4 (30.8) | 4 (44.4) | 2 (50.0) | 15 (100.0) |
| Was able to maintain the 1-meter safety distance at the estimated infection time – no. (%) | 2 (15.4) | 5 (55.6) | 1 (25.0) | 4 (26.7) |

## DISCUSSION

HCWs’ COVID-19 infection rate vs the general population

Overall, 41 or 4.7% of participants had tested positive for COVID-19, which was more than the overall HUS HCWs (0.9%) and much higher than that of general population in HUS-area (0.3%). Healthcare worker’s higher risk for COVID-19 infection was also been reported earlier, and our findings are in line with earlier studies^8 13 21^. The confirmed or likely occupational infections represented 53.7% of participants infection, somewhat higher than that of all HUS-area HCWs (44.0%)^10^. It is notable that the participants also had non-occupational contacts with other HCWs, and in our study, 5 (33%) of confirmed non-occupational infections were still from a colleague.

### FFP2/3 respirator vs Surgical mask in occupational infections

This is the first study that analyses the source of infection and compares the impact of using of either a surgical mask or an FFP2/3 respirators on HCWs’ COVID-19 infections, which alings with an earlier meta-analys indicating that respirators might have a stronger protective effect than surgical masks toward SARS-CoV-2^21^.

In our study, none of the ICU HCWs got sick while using the recommended PPE (FFP3 respirator [FFP2, if FFP3 is not available], pair of gloves, long-sleeved fluid repellent gown, hair protection and eye protection), although they spend the whole shift in the same room with the COVID-19–patients. The OR for occupational-COVID-19 infection was 3.4 (p = 0.016) when working in areas where COVID-19 positive patients are treated. All occupational infections that were traced from patients (N = 4, 20.0%) in the wards happened after instructing HCWs to reduce PPE to surgical masks. In the ICU, the instructions about respirator, PPE and other infection prevention were equal to those recommended in the literature. Although the HUS practice of frequent doffing inside patient area in the wards might increase the risk of infection, the UK guideline is to use the same mask for a session of work, and the infection rate for HCWs is even higher^8 22^. With other viral infections, contamination while D&D has been observed^23^, marking the importance of assisted D&D and training^24^. In our data, only under 40% of participants had trained D&D, and prevention of possible contamination during D&D needs more attention.

In an earlier study, surgical masks were shown to reduce SARS-CoV-2-virus transmission^21^, but in our study, especially in the wards with high exposure, the surgical mask is obviously less protective against COVID-19.

Out of the 413 participants who perform AGPs with infection patients, only 180 (43.6%) use FFP2/3 respirators. Furthermore, 42 (10.7%) participants said that they are not using FFP2/3 respirators even while performing AGPs with COVID-19-positive patients. The WHO and other international and THL recommendations suggest always using FFP2/3 respirators in AGPs^13 19^ ^25^.

Surprisingly, out of 279 participants who performed COVID-19 PCR sampling, 148 (53.0%) used FFP2/3 masks, marking a higher figure than those who care for the confirmed COVID-19 patients. The extensive PCR sampling safety measures, such as drive-in sampling stations where the suspected COVID-19 infected stays in their car for the whole procedure, and the short exposure time of less than 15 minutes during the sampling are likely to explain the absence of COVID-19 patients with laboratory nurses.

### Occupational infections between co-workers

When tracing, 29.3% of infections were traced back to colleagues both at the workplace (N = 7) and outside (N = 5). The use of surgical masks in employees’ facilities could reduce transmissions, but this needs to be studied further. The WHO recommends using medical masks continuously throughout a shift, apart from eating and drinking. The mask should be changed after caring for a patient requiring droplet/contact precautions for other reasons(e.g., influenza) to avoid any possibility of cross-transmission.^13^ At HUS, the guideline has been to use surgical masks only during patient contact, avoid any personal gatherings, and keep at least 1 meter social distance. A recent systematic review^21^ shows a clear association in the reduction of infections by having at least 1 meter of physical distance, and this study also supports the importance of social distances also for HCWs. Despite the restrictions, 83.3% of those who got the infection from a colleague at the workplace and 60.1% of all participants were not able to main the instructed 1-meter radius from other people. Of all participants, 69 were guaranteed due to COVID-19 exposure from a colleague, and 46 of those (66.6%) were not able to maintain the instructed safety distance. This highlights the difficulty of following this instruction in a crowded hospital setting and urges hospitals to find additional ways of reducing transmission between colleagues.

### Other findings

Out of 21 pregnant participants, 10 treated COVID-19 patients, and 3 were uncertain whether they had or not. A recent study from Alzamora et al. showed the possible transmission of the virus from mother to foetus^26^; the consequences of first- or second-trimester infection are still unclear, and considering the hypercoagulopathy of COVID-19 and pregnancy, the risk of thromboembolic events is probably elevated^27^. Luckily, the low risk for perinatal transmission at the time of delivery by non-severe SARS-CoV-2–positive mothers has been reported^28^. More alarming, there have been findings of SARS-CoV-2 in amniotic fluid and placental tissue and maternal death with severe SARS-CoV-2–positive mothers^29 30^. Accordingly, it can be presumed that pregnant women should not engage in direct contact with COVID-19 patients^27 31^.

### Strengths and weaknesses of this study

This is the first study to analyse the impact of using surgical masks vs FFP2/3 respirators in HCWs during the COVID-19 pandemic. The masks and respirators were tested by the employer before usage, and the availability of masks was good during the whole study. The positive COVID-19 infections were proven by COVID-19 RT-PCR and antibody tests, which are the gold standard in COVID-19 diagnostics^32^. The study was conducted prospectively during the COVID-19 pandemic, and all of the nurses and doctors working at HUS were informed about the study. Of those that opted to participate, 95% stated that they have followed the state restrictions. These factors, combined with the low density of 176/km² in Uusimaa, greatly reduce the likelihood of non-occupational infections of HCWs and hence increase the reliability of analysis on workplace-related infections.

It could be argued that the high infection rate of HCWs is due to their more extensive testing. However, the guidelines in Finland have been to test everyone with matching symptoms through universal healthcare. Although the testing HCWs has been prioritised, the testing capacity has been sufficient during most of the first wave, leaving only a limited number of potential symptomatic COVID-19–infected non-HCWs without diagnosis; thus, testing cannot explain the major difference in infection rates.

The characteristics of the participants reflect the overall personnel, although females are slightly overrepresented in this study, and as usual, the people related to the topic participate more frequently, as evident by the number of COVID-19 infected participants. The high number of infected participants gives this study a higher reliability in analysis of infection sources and PPE usage of those infected.

The participants reported that the availability of PPE, masks and respirators has been good compared to the use recommendations, and this does not limit the reliability of our results unlike in many other countries^33^.

## CONCLUSIONS

HCWs have a higher risk of COVID-19 infection compared to the general population. Despite the high exposure to SARS-CoV-2, none of the ICU workers got clear occupational COVID-19 infection, whereas working in a cohorted COVID-19 ward or a normal ward with COVID-19 patients seems to have a high-risk association for occupational COVID-19 infection. The main difference is that the ICU workforce has used FFP2/3 respirators and aerosol precautions, whereas in other departments, the PPE quality has been significantly lower or completely absent. Notably, 29.3% of the infections were from colleagues, thus also requiring special attention for social distances and infection control measures between co-workers. We recommend that PPE similar to what is used in the ICU be used in all COVID-19–related treatments. Per the Finnish Employee Protection Law, this should be done regardless of additional PPE expenses, especially as the global supply of respirators has increased substantially. The possible role of aerosols in the transmission of SARS-CoV-2 needs to be further studied.

## Data Availability

Data sharing: No additional data is available at this point.

## Abbreviations

(COVID-19): coronavirus disease 2019
(PPE): personal protective equipment
(AGP): aerosol-generating procedure

## ACKNOWLEDGEMENTS

We thank Catharina Pomoell for her great work as a research nurse and Johan Sanmark for his support. We also thank all participants involved in this study and their help to find ways to ensure safe working environment.

